# Climatic Drivers of Malaria risk in Children Under Five: A Large-Scale Analysis of individual-level data for 350,000 children in 26 Sub-Saharan African Countries

**DOI:** 10.64898/2026.06.17.26355855

**Authors:** Manuel Martellini O Nocentini, Shivang Pandey, Leonardo Olivetti, Francesco Defilippo, Max Wybrant, Koffi Worou, Maurizio Mazzoleni, Eugenia Quiros-Roldan, Pauline Byakika, Carlo Torti, Cecilia Magnusson, Antonio Gasparrini, Gabriele Messori, Elena Raffetti

## Abstract

**Background:** Malaria risk is influenced by climatic conditions, and children under five are particularly vulnerable due to their limited acquired immunity. We investigate the association between climatic factors and malaria risk in 350,000 children aged 5-59 months in sub-Saharan Africa over 18 years.

**Methods:** We included children aged 5-59 months with malaria tests from Demographic and Health Surveys (DHS) in 26 sub-Saharan African countries between 2006 and 2023. We linked these data to high-resolution climate exposures: temperature, precipitation, soil moisture, actual evapotranspiration and specific humidity. We fitted a mixed-effect logistic regression model incorporating Distributed Lag Non-linear Models (DLNM) over 1-6 month lag window for each exposure, controlling for seasonality and long-term trends. We examined effect modification by maternal education, household wealth, residential type, water source, sanitation facility, child age and sex, use of insecticide-treated bed nets (ITNs), and the age of the household head.

**Results:** Malaria prevalence was 19.5%. Malaria risk was highest at 24 °C (OR: 1.45, 95% CI: [1.36, 1.54]), followed by a decline at higher temperatures. This elevated risk was mainly driven by short-term exposures (1-2 months). Precipitation increased risk up to ~ 120 mm (1.10, [1.07, 1.12]), after which heavier rainfall reduced risk, particularly at short-to medium-term lags (1-4 months). Soil moisture was associated with increasing risk up to ~80 mm (1.11, [1.08, 1.14]), with a plateau at higher levels. Evapotranspiration showed a strong, near-linear positive association with malaria risk. Higher specific humidity levels (>14 g/kg) presented a lower risk, reaching a 45% reduction at 17 g/kg (0.55, [0.49, 0.61]), with the strongest protective effects at short-term lags (1-2 months). Elevated malaria risk at low and moderate average temperatures was particularly evident among children who did not sleep under an ITN net.

**Conclusion:** Malaria risk in children under five is strongly shaped by climatic factors, with complex and delayed associations. The findings provide evidence to guide targeted interventions and early-warning strategies for vulnerable populations.

## 1. Introduction

Malaria is a vector-borne disease transmitted by *Anopheles mosquitoes*. It remains a major global public health challenge, with populations in sub-Saharan Africa carrying a disproportionate burden. According to the latest World Health Organization Malaria Report [1], there were an estimated 263 million cases worldwide in 2023, resulting in 597,000 deaths. This is 11 million more cases than 2022, underscoring persistence of malaria despite ongoing control efforts. Approximately 95% of deaths occurred in the WHO African Region, highlighting the region’s disproportionate burden.

Children under five years of age face the highest malaria risk. As further reported in the 2024 World Malaria Report, they accounted for 76% of all malaria deaths in 2023 in the WHO African Region. This vulnerability stems from the limited acquired immunity and immature immune systems [2-3].

*Anopheles mosquitoes*, the primary vectors of *Plasmodium* parasites, are highly sensitive to environmental conditions [4]. Moderately hot temperatures can accelerate the development of both *Anopheles mosquitoes* and *Plasmodium*, increasing transmission potential [5], while extreme heat may reduce mosquito survival and fecundity, thereby limiting transmission [6]. Precipitation shapes breeding habitats: moderate rainfall supports larval development, whereas intense rainfall can wash away breeding sites and reduce transmission [7]. High humidity improves mosquito survival and activity [8]. High soil moisture sustains larval habitats and supports vector densities [9]. Evapotranspiration, though less studied, can influence breeding site persistence and transmission dynamics [10].

Research across sub-Saharan Africa has linked climatic and environmental factors to malaria risk (e.g. [11-14]). Few studies, however, focus specifically on children under five [15-22], the group at highest risk of severe disease and death. No large-scale analysis using individual-level data has been conducted. Existing work is often constrained by small sample sizes [15, 16, 17, 18, 20] or by reliance on aggregated data [19, 21, 22], and most analyses are limited to single countries or regions (Nigeria [15], Burkina Faso [16, 18], Ghana [17, 20, 22], Western Kenya [19] and Togo [21]). Additionally, environmental effects on health are non-linear and delayed [23], meaning that outcome can be influenced by both present and past environmental exposures. Such lagged dynamics are critical to malaria transmission [24] but are often overlooked.

To address these challenges, this study aims to advance the understanding of the pattern of association between climatic factors and malaria risk in children under five across 26 sub-Saharan countries accounting for complex exposure-response and temporal relationships. To this end, we used individual-level malaria test data from approximately 350,000 children collected over a 17-year period linked with temperature, precipitation, humidity, soil moisture and evapotranspiration data.

## 2. Methodology

### 2.1 Data source and sampling procedure

We used data from the Demographic and Health Surveys (DHS) and Malaria Indicator Surveys (MIS) conducted across 26 countries in sub-Saharan Africa from 2006 to 2023: Angola, Burkina Faso, Benin, Burundi, Democratic Republic of the Congo (DRC), Côte d’Ivoire, Cameroon, Gabon, Ghana, Gambia, Guinea, Kenya, Liberia, Madagascar, Mali, Mauritania, Malawi, Mozambique, Nigeria, Niger, Rwanda, Sierra Leone, Senegal, Togo, Tanzania and Uganda.

Both DHS and MIS surveys were implemented by the DHS Program, using standardised questionnaires and a common sampling methodology; we therefore refer to them collectively as “DHS” throughout this paper [25].

The DHS is a nationally representative household survey that collects data on key health indicators, including malaria. A stratified two-stage cluster sampling was applied in each country. First, the country was divided into administrative units, referred to as Enumeration Areas or clusters, which are then stratified by region and urban/rural classification. Within the selected cluster, systematic sampling was used to identify the households to be surveyed. This hierarchical structure (children nested within clusters, which are nested within countries) was explicitly accounted for in the modelling approach using a mixed effect model. A detailed description of the sampling procedure is available in the DHS sampling and household listing manual [26]. Survey data included information on household characteristics, such as demographic, socio-economic and health-related information for all eligible household members.

The DHS tested children aged 5-59 months for malaria, regardless of symptoms, using microscopy, Rapid Diagnostic Tests, or both. This age range was selected given maternal antibodies in newborns and the logistical challenges of testing school-aged children. Positive test results were not differentiated by Plasmodium species.

### 2.2 Variables of the study

#### 2.2.1 Climatic Exposures

The DHS survey data contained geo-referenced clusters, with GPS coordinates included for the centroid of each sampled cluster. To ensure confidentiality, DHS applied spatial displacement: 0-2 km for urban clusters, 0-5 km for rural clusters, and up to 10 km for every 100th cluster [27]. We then used these latitude and longitude coordinates to link climatic and environmental exposures to each cluster and examine their association with malaria.

We extracted data from the TerraClimate dataset [28] at a spatial resolution of ~4.6 km andmonthly resolution for minimum and maximum temperatures (°C), precipitation accumulation (mm), soil moisture (mm) and actual evapotranspiration (mm). Minimum and maximum temperatures were averaged to derive a single average temperature variable. Specific humidity (g/kg) was sourced from the Famine Early Warning Systems Network (FEWS NET) Land Data Assimilation System (FLDAS) dataset at spatial resolution of ~11.1 km [29], which was rescaled to ~5 km before extraction using bilinear interpolation. Bilinear interpolation computes a distance-weighted average along the x- and y-axis, yielding a smooth, continuous field [30].

Climatic variables were extracted for lags of 1 to 6 months before the month of malaria testing [31-32], in order to capture short- and medium-term effects.

#### 2.2.2 Outcome Variable

The outcome was malaria infection, coded as binary variable, with 1 indicating presence and 0 indicating absence of infection. When both microscopy and a Rapid Diagnostic Test were available, we used microscopy given its higher reliability [33]. If a microscopy test was missing, but a Rapid Diagnostic Test was available, this was used instead (27.16% of the sample). In the DHS data, the Rapid Diagnostic Test showed a specificity of 87% and a sensitivity of 82% compared with microscopy. We conducted a sensitivity analysis including only children tested by microscopy.

#### 2.2.3 Socio-economic covariates

Based on previous literature, we considered the following socio-economic covariates: sex of the child (Female/Male), age of the child (months), mother’s educational level (No education/Primary-Middle/Secondary-Higher/Missing), use of Insecticide-Treated bed Nets (ITN) the night before the interview (No/Yes), type of residency (Rural/Urban), household source of drinking water (Unimproved/Improved), household type of toilet facility (Unimproved/Improved), age of household head (years) and the Relative Wealth Index (RWI).

All the covariates were derived from the DHS survey, except for RWI and type of residency. The RWI is a proxy for socioeconomic status at the local level derived from satellite imagery and de-identified Facebook connectivity data [34]. It provides a static estimate of relative wealth at ~2.4 km resolution using data from 1st April 2021 and 22nd December 2023. Type of residency was derived from the Global Human Settlement Layer (GHSL) dataset [35], which classifies the degree of urbanization at 1 km resolution in 5-year intervals. Table 1 summarises how all the previously mentioned covariates were operationalised.

**Table 1:** Socio-economic covariates and their operationalisation.

| Covariate | Operationalisation |
| --- | --- |
| Sex of the child | The variable sex of the child was categorised into two groups: Female or Male. |
| Age of the Child | The variable age of the child was treated as a continuous variable. |
| Mother's educational level | The variable mother's educational level was categorized in four groups: No education, Primary/Middle, Secondary/Higher or Missing. The "Missing" category was included to retain approximately 40000 that would otherwise be lost. |
| ITN use | The variable ITN use was categorised into two groups: Yes or No. |
| Type of Residency | The variable type of residency was categorised into two groups: Urban or Rural. |
| Household source of drinking water | The variable Household source of drinking water was categorised into two groups: Unimproved or Improved. |
| Household type of toilet facility | The variable household type of toilet facility was categorised into two groups: Unimproved or Improved. |
| Age of head of household | The variable age of head of household was treated as a continuous variable. |
| Relative Wealth Index (RWI) | The variable Relative Wealth Index (RWI) was treated as a continuous variable. |

#### 2.2.4 Confounders

We adjusted for seasonality and long-term trends by including calendar month and survey year as individual-level covariates. Month was modelled with a natural cubic B-spline with four degrees of freedom, and year as a linear term.

We fitted separate models for each climatic exposure. Confounder selection was guided by a Directed Acyclic Graph (DAG; Figure S1) to control for upstream climatic drivers while avoiding over-adjustment for mediators. Temperature and precipitation were mutually adjusted; the soil-moisture model additionally included temperature and precipitation; the evapotranspiration model further adjusted for soil moisture; and the humidity model included all four variables. Details are provided in Supplementary Table S1. For each of these confounding variables, the average value across all lags was computed and included as a covariate in the corresponding model.

### 2.3 Statistical analysis

#### 2.3.1 Statistical model

We used mixed-effects logistic regression models to examine the association between climatic exposure and malaria outcome. We considered the following hierarchical structure: children (level 1) nested within clusters (level 2), which are in turn nested within countries (level 3).

A Distributed Lag Non-linear Model (DLNM) was integrated in the mixed effect model, to capture the non-linear and delayed effects of the exposures. DLNM models are based on the concept of cross-basis, a bi-dimensional function that simultaneously characterizes the exposure-response and lag-response relationships [36]. Two sets of basis functions are applied: one set to the original exposure and another to the lag vector ***ℓ***^*T*^ = (*ℓ*_0_, …,*l*, …,*L*), where *ℓ*_0_ and *L* correspond to the minimum and maximum lag, respectively. This allows to capture both the non-linear exposure-response relationship and the shape of the association across lags.

DLNM models are primarily used in time-series analyses. However, they can also be applied to individual-level data, as in our study. This is done by constructing a matrix of exposure histories, representing the series of exposures experienced at each lag for each child [37].

Climatic variables were considered individual level exposures. This is because some households within the same cluster were surveyed in different months, and 149 clusters were surveyed in two different years.

Let *Y*_*ijk*_ be the binary outcome indicating whether child *i* ∈ {1, …, *n*_*jk*_} nested in cluster *j* ∈ {1, …, *J*_*k*_} which is nested in country *k* ∈ {1, …, *K*} tested positive (1) or negative (0). The model is specified as:

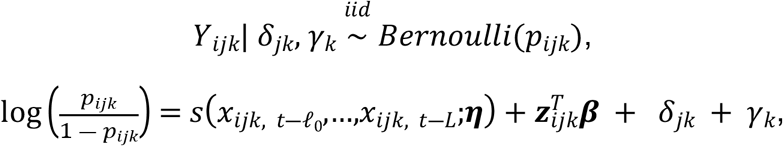

Here *x*_*ijk, t*―*l*_ represents the value of the environmental exposure *x* at lag *l*. The function *s* allows non-linear relationships and delayed effects between the exposure and the outcome. ***z***_*ijk*_ is a vector containing all the variables associated with the fixed effect coefficients ***β***, including the average value across lags of the controlled environmental exposures, month (as a spline), and year (as a linear term). Since the climatic variables were extracted from one to six months prior to the month of testing, *ℓ*_0_ = 1 and 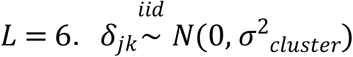 are the cluster random effects and 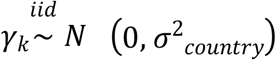 are the country random effects, assumed mutually independent.

The function 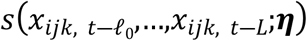 is based on the concept of cross-basis explained above. Let 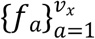 be the basis for the exposure and 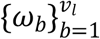 the basis for the lags.Then:

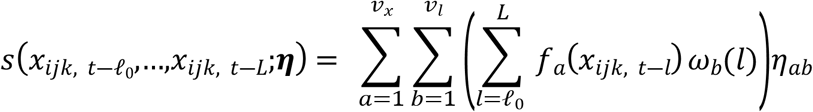

Following Gasparrini et al. [38], this term can be expressed in matrix notation:

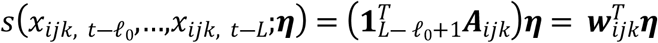

where ***w***_*ijk*_ is obtained by summing along the columns of the (*L* ― *ℓ*_0_ +1) × *v*_*x*_ · *v*_*l*_ matrix ***A***_*ijk*_. Define ***R***_*ijk*_ as the (*L* ― *ℓ*_0_ +1) × *v*_*x*_ matrix of basis transformations applied to the vector 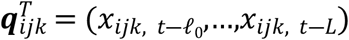. Let ***C*** be the (*L* ― *ℓ*_0_ +1) × *v*_*l*_ matrix of basis variables applied to the lag vector. The matrix ***A***_*ijk*_ can be then obtained as:

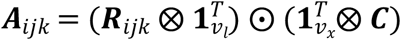

where ⨂ and ⨀ represent the Kronecker and the Adamard product, respectively. The cross-basis matrix ***W*** is then obtained by applying the previous calculations to all *i* · *j* · *k* observations and can then be used as a model matrix in the model.

For each climatic exposure, the exposure-response relationship was modelled with a natural cubic B-spline with knots at the 33.3rd and 66.6th percentiles, treating all lags jointly as a single vector. The lag-response relationship was modelled with a natural cubic B-spline with a knot at 3.5 months. These basis specifications provide sufficient flexibility to capture both associations.

After the parameters ***η*** have been estimated, the effect at a specific exposure level *x*_*p*_ and lag *l*_*p*_ can be computed as:

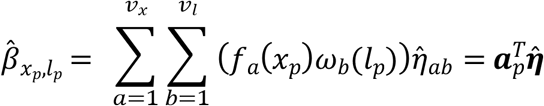

Here, ***a***_*p*_ is a *v*_*x*_ · *v*_*l*_ × 1 vector, defined analogously to ***A***_*ijk*_,but using vectors instead of matrices. Let ***r***_*p*_ and ***c***_*p*_ be the *v*_*x*_ × 1 and *v*_*l*_ × 1 vectors obtained by applying the basis transformation to *x*_*p*_ and *l*_*p*_, respectively. Then:

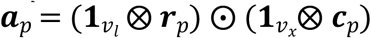

This formulation allows estimation of effects across a grid of exposure and lag values, producing a 3D surface that represents the exposure-lag-response association. Additionally, two-dimensional plots can be obtained by fixing one argument: plotting effects against exposure, or against lags at selected exposures.

The variance of 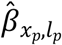 is derived from the variance-covariance matrix of 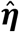:

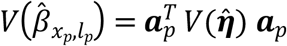

It is also possible to estimate the cumulative effect of an exposure *x*_*p*_, by summing all the effects across all lag values ***ℓ***. Assuming *x*_*p*_ remains fixed across the lag period:

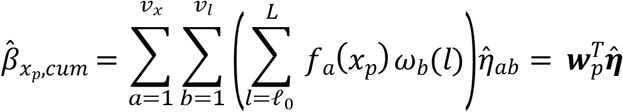

Here, ***w***_*p*_ is defined in the same way as ***w***_*ijk*_, but constructed using the fixed exposure value *x*_*p*_ across all lags. The variance of the cumulative effect is:

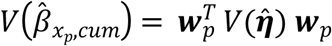

For interpretability, effects were centred at a reference exposure (the median across all lags) and exponentiated to yield odds ratios. We evaluated effects over a grid from the 2.5th to 97.5th exposure percentiles to reduce the influence of extremes on the curve tails.

All socio-economic covariates were modelled at the individual-level, except RWI and residency type, which were considered cluster-level covariates. RWI is time-invariant and residence type is available only at five-year intervals. We first fitted a mixed-effects logistic model including all covariates to estimate main effects while controlling for the mean across lags of all climatic variables. Individuals with any missing covariate were excluded. We then assessed effect modification by fitting separate models with interactions between the cross-basis and each covariate. Estimation details for interaction models are provided in the Supplementary Methods.

We also conducted a series of sensitivity analyses. These included restricting the analysis to microscopy-tested children and adjusting for individual lagged variables, instead of using lag averages for confounders.

All data management and statistical analysis were performed using R software 4.4.2. Models were fitted with the `glmmTMB()`function from the `glmmTMB` package [39-40]. Further details are given in the Supplementary Methods.

## 3. Results

### 3.1 Characteristics of the sample

The analysis included data from 26 sub-Saharan African countries from 2006 to 2023, with an initial sample of 357,085 children. Supplementary table S2 presents the distribution of children aged 6-59 months by country and year. We excluded 4,057 children with missing GPS data, 4,436 with inconclusive malaria tests, and 27 with both, yielding a final sample of 348,565 children from 25,813 clusters.

Table 2 summarizes the socio-demographic characteristics. Overall malaria prevalence was 19.5%. The median age was 32 months, and 50.6% were male. About 41% of mothers had no formal education, while nearly 20% had completed secondary or higher education. Over half (55.4%) of the children slept under an insecticide-treated net the night before the survey and 59.7% lived in rural areas. Most children (68.8%) had access to improved drinking water, but only 42.9% had improved sanitation. Household heads had a median age of 41 years.

**Table 2:** Descriptive Statistics of the variables of the sample.

| Variable | Missing values (%) | Categories | Frequency (%) |
| --- | --- | --- | --- |
| Test result | 0 (0.00) | Negative | 280704 (80.53) |
|  |  | Positive | 67861 (19.47) |
| Sex of the child | 0 (0.00) | Male | 176253 (50.57) |
|  |  | Female | 172312 (49.43) |
| Mother's educational level | 0 (0.00) | No education | 141925 (40.72) |
|  |  | Primary/Middle | 100660 (28.88) |
|  |  | Secondary/Higher | 64984 (18.64) |
|  |  | Missing | 40996 (11.76) |
| ITN net | 0 (0.00) | No | 155426 (44.59) |
|  |  | Yes | 193139 (55.41) |
| Residency | 1027 (0.29) | Rural | 208065 (59.69) |
|  |  | Urban | 139473 (40.02) |
| Household source of drinking water | 45 (0.01) | Unimproved | 108877 (31.24) |
|  |  | Improved | 239643 (68.75) |
| Household type of toilet facility | 79 (0.02) | Unimproved | 198805 (57.04) |
|  |  | Improved | 149681 (42.94) |
|  |  | Min, Max | Median (IQR) |
| Age of the child (months) | 0 (0.00) | 5, 59 | 32 (19, 46) |
| Age of the head of the household (years) | 1184 (0.40) | 10.00, 97.00 | 41.00 (33.00, 53.00) |
| Relative Wealth Index (RWI) | 0 (0.00) | -1.45, 2.43 | -0.23 (-0.49, 0.35) |
| Average Temperature (lag 1) (°C) | 0 (0.00) | 8.90, 35.50 | 26.05 (23.75, 27.45) |
| Precipitation Accumulation (lag 1) (mm) | 0 (0.00) | 0.00, 1782.00 | 90.00 (17.00, 194.00) |
| Soil Moisture (lag 1) (mm) | 0 (0.00) | 0.00, 556.40 | 80.40 (24.60, 181.00) |
| Actual Evapotranspiration (lag 1) (mm) | 0 (0.00) | 0.00, 184.30 | 90.20 (46.80, 111.40) |
| Specific Humidity (lag 1) (g/kg) | 0 (0.00) | 2.12, 20.00 | 14.28 (11.46, 16.39) |

Table 2 and Figure 1 present descriptive statistics and distributions of the climatic exposures at lag 1.

**Figure 1.**
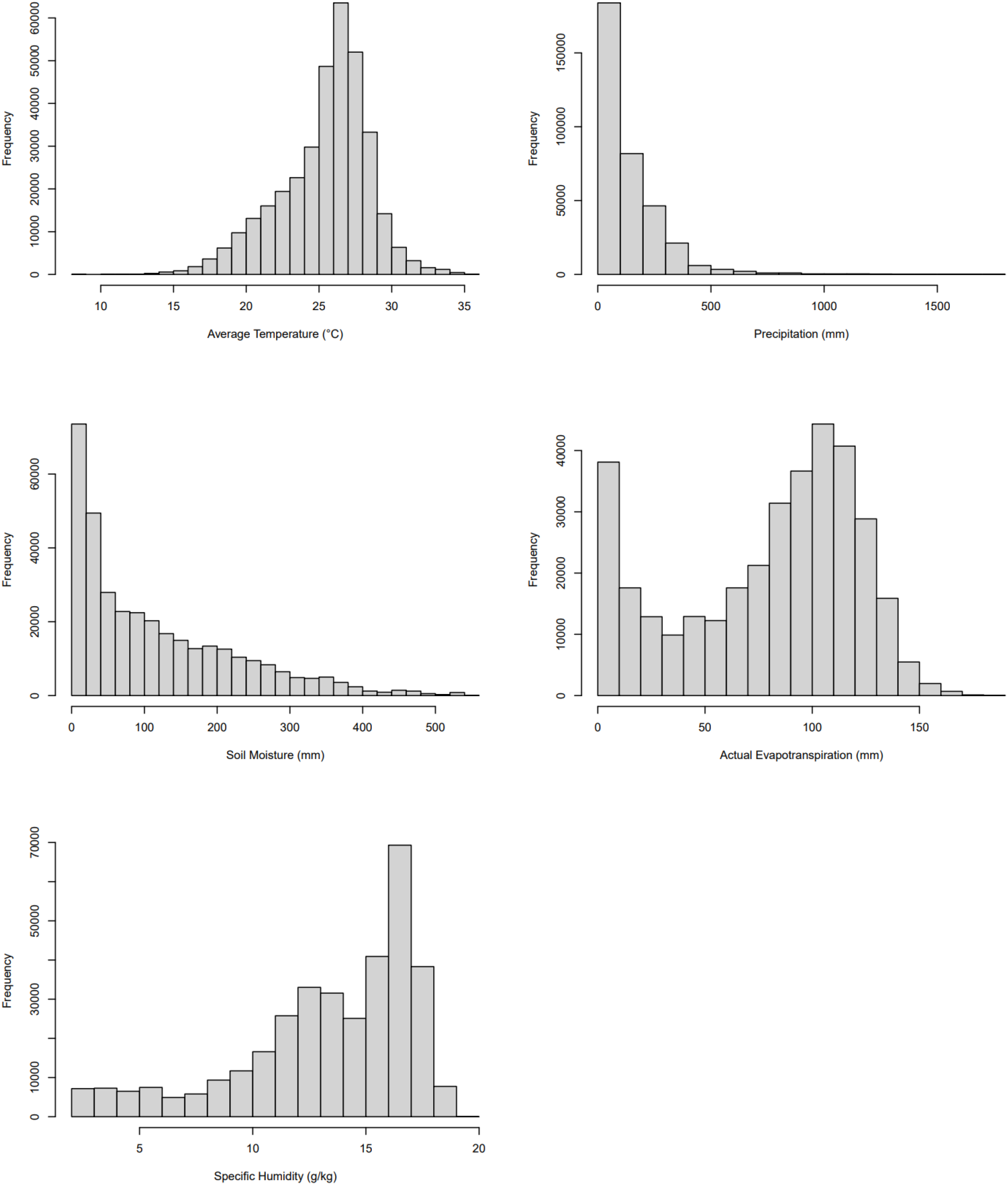
Distribution of climatic variables at lag 1.

### 3.2 Models Results: Associations between Climatic Factors and Malaria

#### 3.2.1 Cumulative associations

Figure 2 shows the overall cumulative effects of climatic exposures on malaria risk over 6 months (95% confidence intervals, CIs, in brackets). For temperature, malaria risk peaked at 24°C, corresponding to a 45% increase compared to the reference 26°C (CI: [1.36, 1.54]). Risk declined at both lower and higher temperatures and flattened above 30 °C. For precipitation, risk increased up to 120 mm, at which point odds of being infected were 7% higher compared to the reference level of 86 mm (CI: [1.07, 1.12]). A protective effect was observed at higher exposure levels, though not significant. Soil moisture was associated with increasing malaria risk up to ~80 mm, where the risk was 11% significantly higher, compared to the reference level of 57 mm (CI: [1.08, 1.14]). The effect then flattened and became not significant. Actual evapotranspiration displayed a strong near-linear positive association with malaria risk, with risk 63% higher at 120 mm compared to 88 mm (CI: [1.48, 1.80]). In contrast, specific humidity was protective at higher levels. Above ~15 g/kg, risk declined, reaching a 45% reduction at 17g/kg, relative to the reference level of 14 g/kg (CI: [0.49, 0.61]).

**Figure 2.**
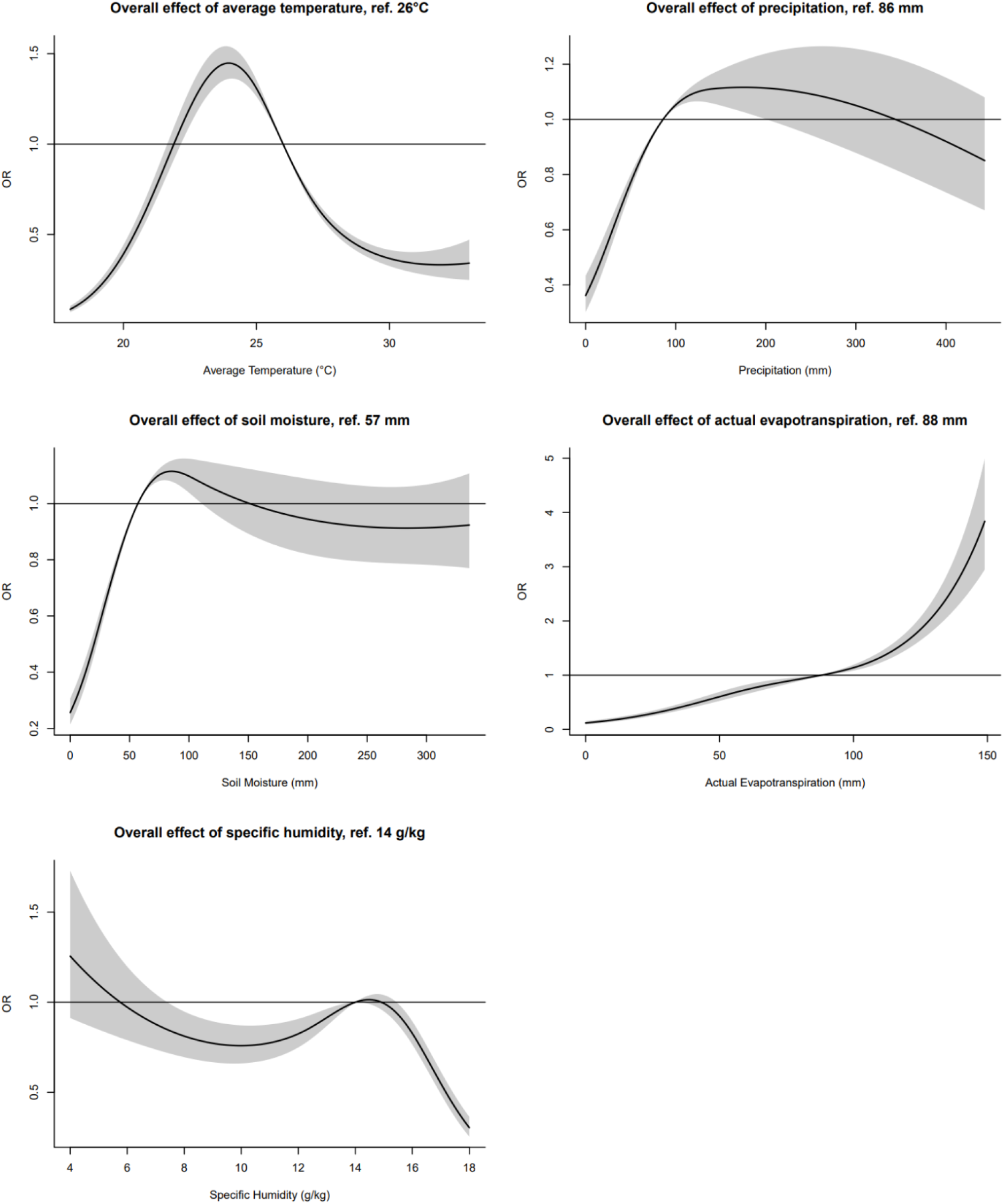
Cumulative effect of climatic exposures on malaria risk at lags 1-6 months.

Results remained consistent in sensitivity analyses (Figures S2-S3). Notably, in the microscopy-based analysis, high-temperature effects were more pronounced.

#### 3.2.2 Associations along lags

Figures 3 shows the exposure-lag-response and lag-response associations between climatic exposures and malaria risk. For average temperature (Figure S4), the elevated risk observed around 24°C was primarily driven by short-term exposures (lags 1-2). Although higher temperatures generally had a protective effect, a delayed increase in risk appeared at lag 6. For precipitation (Figure S5), protective effects at higher precipitation (>150 mm) emerged at short-medium lags, particularly between lags 1-4. However, at longer lags, risk increased again with 400 mm at lag 6 associated. The elevated risk around 120 mm was largely attributable to longer lags (5-6). Soil moisture (Figure S6) showed increased risk around 80-200 mm during early lags (1-2), while higher moisture levels were associated with a higher effect at medium-term lags (2-4). Actual evapotranspiration (Figure S7) generally displayed a positive association with malaria risk at high levels (>100 mm), particularly at lags 1 and 6. Specific humidity (Figure S8) was associated with reduced malaria risk at high levels (>14 g/kg), especially during early lags (1-2).

**Figure 3.**
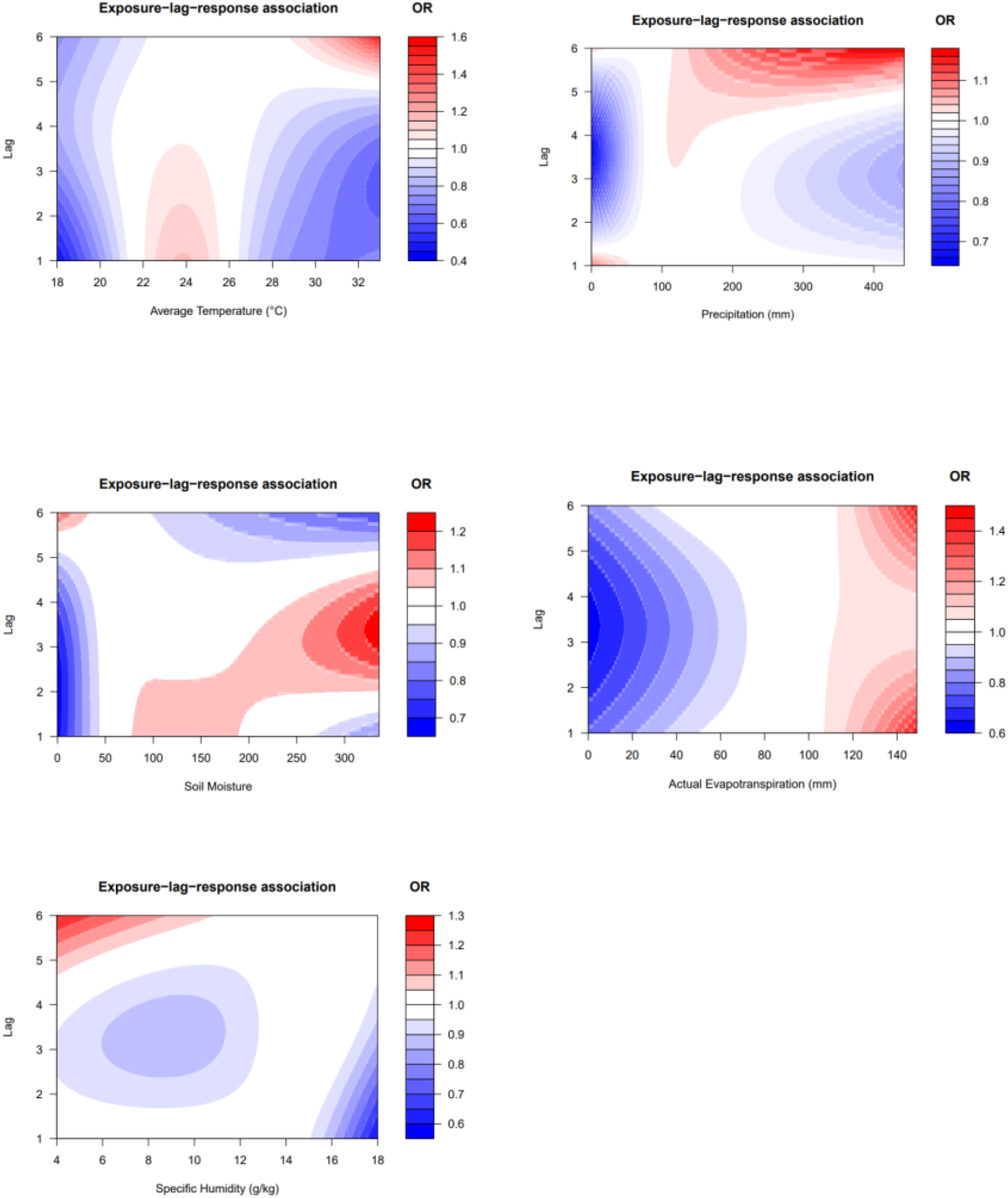
Contour plots of exposure-lag-response relationship between climatic exposures and malaria risk at lags 1-6 months.

#### 3.2.3 Socio-economic covariates and effect modification

We then focused on the association between socio-economic variables and risk of malaria controlling for the average across lags of all climatic variables. Higher Relative Wealth Index (OR = 0.36, CI: [0.35, 0.38]), higher maternal education (primary/middle: OR = 0.88, CI: [0.86, 0.91]; secondary/higher: OR = 0.59, CI: [0.57, 0.62]), ITN use (OR = 0.84, CI: [0.82, 0.86]), improved toilet facilities (OR = 0.76, CI: [0.74, 0.78]), improved source of drinking water (OR = 0.83, CI: [0.80, 0.85]), and urban residence (OR = 0.94, CI: [0.89, 0.98]) reduce the risk. Being male (OR = 1.03, CI: [1.01, 1.05]), increasing age of the child (OR = 1.021, CI: [1.020, 1.022]) and household head (OR = 1.001, CI: [1.000, 1.002]) were associated with slightly higher risk. In the interaction analyses (Figures S9-S13, Table S4), most socio-economic covariates showed statistically significant interactions with climatic variables. However, these effects were generally small compared to the main effects, and thus few were of clinical relevance. Particularly, risk was higher at low and moderate average temperatures among children who did not sleep under an ITN, and very low humidity levels appeared to be more protective for low wealth index values.

## 4. Discussion

In this study, we characterised the associations between climatic variables and malaria risk in children aged 6-59 months in 26 sub-Saharan African countries. The main findings were that malaria risk followed an inverted U-shaped relationship with temperature, peaking at moderate levels; increased with precipitation and soil moisture up to moderate values before plateauing; increased almost linearly with evapotranspiration; and declined at high levels of humidity.

We identified a thermal threshold for malaria risk: risk increased up to ~24°C, with elevated risk at short-term lags (1-2 months), then declined and plateaued at higher temperatures. Notably, at very high temperatures, a pronounced effect re-emerged at 6-month lag. These findings are consistent with evidence showing a transmission peak around 25°C, followed by a decline at warmer conditions [6]. The elevated short-term risk likely reflects the Extrinsic Incubation Time of 10–15 days near this optimal temperature [41, 42]. The delayed effect at high temperatures may occur because, although high temperatures accelerate mosquito development [41, 42], the resulting adults are often smaller, less fit, and predominantly male, which delay transmission [43].

Malaria risk increased with precipitation at low to moderate levels, with a peak around 120 mm at longer lags (5-6). Heavier precipitation levels were associated with lower risk at short-medium lags (1-4), but higher risk at longer lags (5-6), suggesting that heavy rain can have lasting effects [44]. Prior evidence, even if focusing on a specific region of South Africa, indicates an optimal monthly rainfall range for malaria transmission of 95-125 mm [45], in line with our analysis. The short-term protective effect observed at high levels of precipitations is consistent with intense rainfall washing away breeding sites, reducing transmission [7, 46], while the long-term risk may result from permanent or semi-permanent water bodies, after rainfall, which sustain *Anopheles* breeding [47].

Soil moisture showed a similar pattern. Higher risk was observed around 80 mm at short-term lags (1-2 months), flattening at higher levels. At medium-term lags (2-4), higher soil moisture was again associated with increased risk. Given the close link between soil moisture and precipitation, these patterns support that the increase in soil moisture implies an increase in oviposition by females [48], whereas excessive soil saturation may reflect temporary flooding or unstable habitats that are unsuitable for mosquito survival. The increased risk associated with higher soil moisture levels is likely related to the fact that the increased soil water storage following repeated rainfall events can prolong breeding sites and enhances mosquito survival [49]. This is consistent with the longer-lag effects we observed for heavy precipitation.

The strongest association was observed for actual evapotranspiration: malaria risk exhibited a strong and positive association, consistently increasing across the entire range of the variable. In particular, we found that the effect of high evapotranspiration levels was pronounced at both longer and, especially, shorter lags. This finding is consistent with previous studies reporting a significant positive association between evapotranspiration and malaria incidence at short-term lags [10]. While the role of evapotranspiration has rarely been investigated in the malaria literature, particularly regarding its lagged effects, our results highlight its importance as a climatic indicator of transmission.

Specific humidity showed an increase in malaria risk around 15 g/kg, beyond which higher humidity levels were associated with a marked reduction in risk, particularly at short-term lags (1-2). This pattern likely reflects the role of humidity in reducing mosquito desiccation and increasing survival up to an optimal threshold, beyond which further increases in humidity may adversely affect mosquito survival, thus affecting transmission. This observation aligns with previous findings [31]. Additionally, the protective effect of higher humidity observed at shorter lag periods, also documented in the same study [31], indicates that the immediate impact of high humidity on mosquito survival play a crucial role in short-term transmission dynamics.

We observed protective associations with malaria risk for several well-established covariates, including higher relative wealth, maternal education, improved household sanitation (toilet facilities and drinking water sources), use of insecticide-treated nets (ITNs), and urban residence [50-53]. In contrast, we found statistically significant increases in malaria risk associated with male sex, older child age, and older age of the household head. However, the magnitude of these effects was minimal (odds ratios ranging from 1.01 to 1.03), thus limiting their potential clinical relevance.

In the effect modification analysis, nearly all socio-economic covariates showed highly significant interactions with the climatic variables, though the overall exposure-response curves were similar across groups. This reflects the complex nature of DLNM models: the interaction test really compares the entire exposure-response-lag surface between groups. With a large dataset, even small differences in these three-dimensional relationships can yield highly significant results. Only few of these interactions were of clinical importance. In particular, the protective effect of ITN use was more pronounced at low and moderate temperature levels, plausibly because higher temperatures reduce ITN use, as bed nets restrict airflow and make sleeping uncomfortable [54], resulting in a smaller effect at higher temperatures. While higher wealth was generally protective, we observed an increased malaria risk at lower humidity levels. This may be due to the very few malaria cases in this region at low humidity levels, driving high levels of uncertainty in the effect estimates at the edges of the distribution.

Our study is based on a large and geographically diverse dataset, approximately 350,000 children under five tested for malaria across 26 sub-Saharan African countries over 17 years. This addresses limitations of prior work that used small samples or single-setting, aggregated data, and improves generalisability. To avoid over-adjustment, we used a Directed Acyclic Graph (DAG) to identify and control for confounding. To improve exposure assessment, we linked high-resolution environmental data (~5 km). To better capture temporal dynamics, we modelled lagged effects using a DLNM model, revealing risk patterns that emerged months after exposure, patterns a traditional cross-sectional approach would miss. To account for the hierarchical clustering of the data, we integrated the DLNM model within a multilevel logistic regression framework. Findings were robust across sensitivity analyses.

This study has some potential limitations. First, the displacement of DHS cluster coordinates to protect confidentiality may have introduced exposure misclassification by shifting clusters into incorrect grid cells. Second, because few children were exposed at the extreme values of some climatic factors, estimates in those ranges are imprecise, with wide confidence intervals and greater uncertainty. Third, the ITN indicator captures only whether the child slept under a net the night before the survey. It does not reflect routine use and may be influenced by symptom-driven use, introducing potential reverse causation. Fourth, we lacked data on vaccination and chemoprophylaxis (including prophylaxis for high-risk groups such as children with sickle cell disease), so residual confounding may remain. If coverage is greatest in high-risk, hot or rainy settings, the bias could attenuate climate-malaria associations. Finally, the Relative Wealth Index (RWI) was derived from 2021-2023 data and may not accurately represent socioeconomic conditions earlier in the study period.

## 5. Conclusion

In this study, we examined the association between climatic variables and malaria risk in children aged 6-59 months using individual-level data from across sub-Saharan Africa. Malaria risk peaked at 24 °C, driven by short-term exposures, and then declined at higher temperatures. Risk increased with precipitation up to around 120 mm, but heavier rainfall levels were associated with reduced risk, particularly over short-to medium-term lags. Soil moisture raised risk up to about 80 mm before flattening out, while evapotranspiration showed a strong positive association with malaria risk. High specific humidity presented lower risk, with the strongest protective effects at short-term lags. Higher risk at low and moderate temperatures was most pronounced among children without ITN use. Our methodology and findings could guide early-warning systems and targeted malaria-control strategies for vulnerable populations.

## Acknowledgements

This research was funded by Formas the Swedish Research Council for Sustainable Development (Formas grant nos. 2022-01845 and 2023-01774) and the Swedish Research Council for Health, Working Life and Welfare (FORTE grant nos. 2024-00833).

M. M. O N.’s work is supported by the Swedish Research Council for Health, Working Life and Welfare (FORTE grant nos. 2024-00833) and the Swedish Research Council for Sustainable Development (Formas grant nos. 2022-01845). S. P.’s work is supported by the Swedish Research Council for Sustainable Development (Formas grant nos. 2023-01774). E. R.’s work is supported by the Swedish Research Council for Health, Working Life and Welfare (FORTE grant nos. 2022-00882 and 2024-00833); the Swedish Research Council for Sustainable Development (Formas grant nos. 2023-01774 and 2022-01845); and the Swedish Research Council (VR, grant nos. 2023-01982 and 2022-06599).

The computations and data handling were enabled by resources provided by the National Academic Infrastructure for Supercomputing in Sweden (NAISS), partially funded by the Swedish Research Council through Grant Agreement No. 2022-06725.

## Data availability

The complete R code used in this analysis is publicly available in a GitHub repository (https://github.com/manumarte00/malaria_climate), which also includes a simulated dataset which resembles the structure of the original DHS dataset.

## Declaration of generative AI and AI-assisted technologies in the writing process

During the preparation of this work the authors used ChatGPT (OpenAI) for grammar and language editing. After using this tool/service, the authors reviewed and edited the content as needed and take full responsibility for the content of the publication. Finally, no AI tools contributed to the scientific content, analysis, or interpretation.

