## Supplementary Material for "Climatic Drivers of Malaria risk in Children Under Five: A Large-Scale Analysis of individual-level data for 350,000 children in 26 Sub-Saharan African Countries"

### Supplementary Methods

#### Interactions

To assess the role of each socio-economic covariate, we included an interaction term between the cross-basis matrix and the covariate into the model. As shown in Table 1, the covariates included both categorical variables (binary or with more than two categories) and continuous variables.
For binary covariates coded as 0 (reference category) and 1 (comparison category), the estimated coefficients from the "main effect" cross-basis matrix, denoted by $\hat{\boldsymbol{\eta}}_{\mathbf{0}}$, represented the exposure-lag-response association for the reference category. To obtain the corresponding effect for the category coded as 1, we summed the "main effect" coefficients $\hat{\boldsymbol{\eta}}_{\mathbf{0}}$ and interaction term coefficients $\hat{\boldsymbol{\eta}}_{\boldsymbol{int}}$:

$$\hat{\boldsymbol{\eta}}_{\mathbf{1}} = \hat{\boldsymbol{\eta}}_{\mathbf{0}} + \hat{\boldsymbol{\eta}}_{\boldsymbol{int}}$$

The variance-covariance matrix for $\hat{\boldsymbol{\eta}}_{\mathbf{1}}$ was computed as:

$$V(\hat{\boldsymbol{\eta}}_{\mathbf{1}}) = V(\hat{\boldsymbol{\eta}}_{\mathbf{0}}) + V(\hat{\boldsymbol{\eta}}_{\boldsymbol{int}}) + V(\hat{\boldsymbol{\eta}}_{\mathbf{0}}, \hat{\boldsymbol{\eta}}_{\boldsymbol{int}}) + V(\hat{\boldsymbol{\eta}}_{\boldsymbol{int}}, \hat{\boldsymbol{\eta}}_{\mathbf{0}})$$

All components were extracted from the full model's variance-covariance matrix $V(\hat{\boldsymbol{\eta}})$.

The approach described above was readily extended to categorical covariates with more than two levels.

For continuous covariates, we obtained the coefficients representing the exposure-lag-response association at a specific value $z$ of the covariate as:

$$\hat{\boldsymbol{\eta}}_{\boldsymbol{z}} = \hat{\boldsymbol{\eta}}_{\mathbf{0}} + z \cdot \hat{\boldsymbol{\eta}}_{\boldsymbol{int}}$$

Here, $\hat{\boldsymbol{\eta}}_{\mathbf{0}}$ corresponds the vector of coefficients from the "main effect" cross-basis, which also represents the effect when the covariate is equal to 0, and $\hat{\boldsymbol{\eta}}_{\boldsymbol{int}}$ corresponds to the interaction term coefficients.
The corresponding variance-covariance matrix for $\hat{\boldsymbol{\eta}}_{\boldsymbol{z}}$ was computed as:

$$V(\hat{\boldsymbol{\eta}}_{\boldsymbol{z}}) = V(\hat{\boldsymbol{\eta}}_{\mathbf{0}}) + z^2 \cdot V(\hat{\boldsymbol{\eta}}_{\boldsymbol{int}}) + z \cdot (V(\hat{\boldsymbol{\eta}}_{\mathbf{0}}, \hat{\boldsymbol{\eta}}_{\boldsymbol{int}}) + V(\hat{\boldsymbol{\eta}}_{\boldsymbol{int}}, \hat{\boldsymbol{\eta}}_{\mathbf{0}}))$$

The effects for continuous covariates were evaluated at the first and third quartile.

Finally, the significance of each interaction terms was assessed using the Likelihood Ratio Test (LRT).

#### Package `glmmTMB`

Like `lme4` package, commonly used for generalised linear mixed-models (GLMMs), `glmmTMB` uses maximum likelihood estimation with Laplace approximation to integrate out random effects. However, `glmmTMB` offers substantial computational advantage, particularly for non-Gaussian models, by leveraging Template Model Builder (TMB). TMB incorporates C++ and automatic differentiation to compute model gradients, enabling faster and more flexible estimation, compared to the `glmer()` function in `lme4`.

### Supplementary Figures

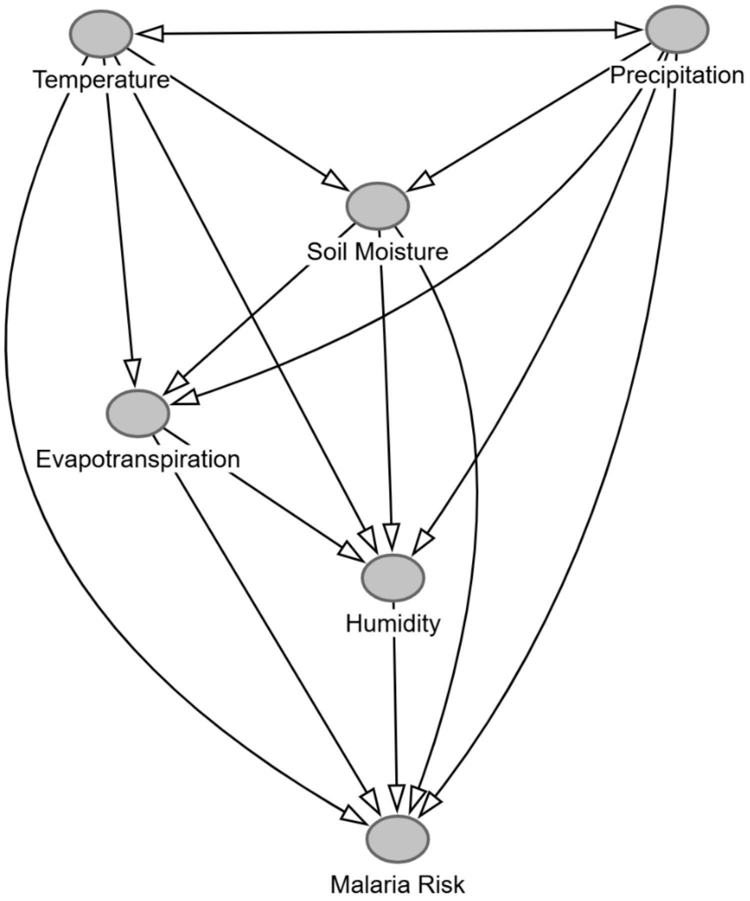

*Figure S1: DAG Graph for climatic exposures.*

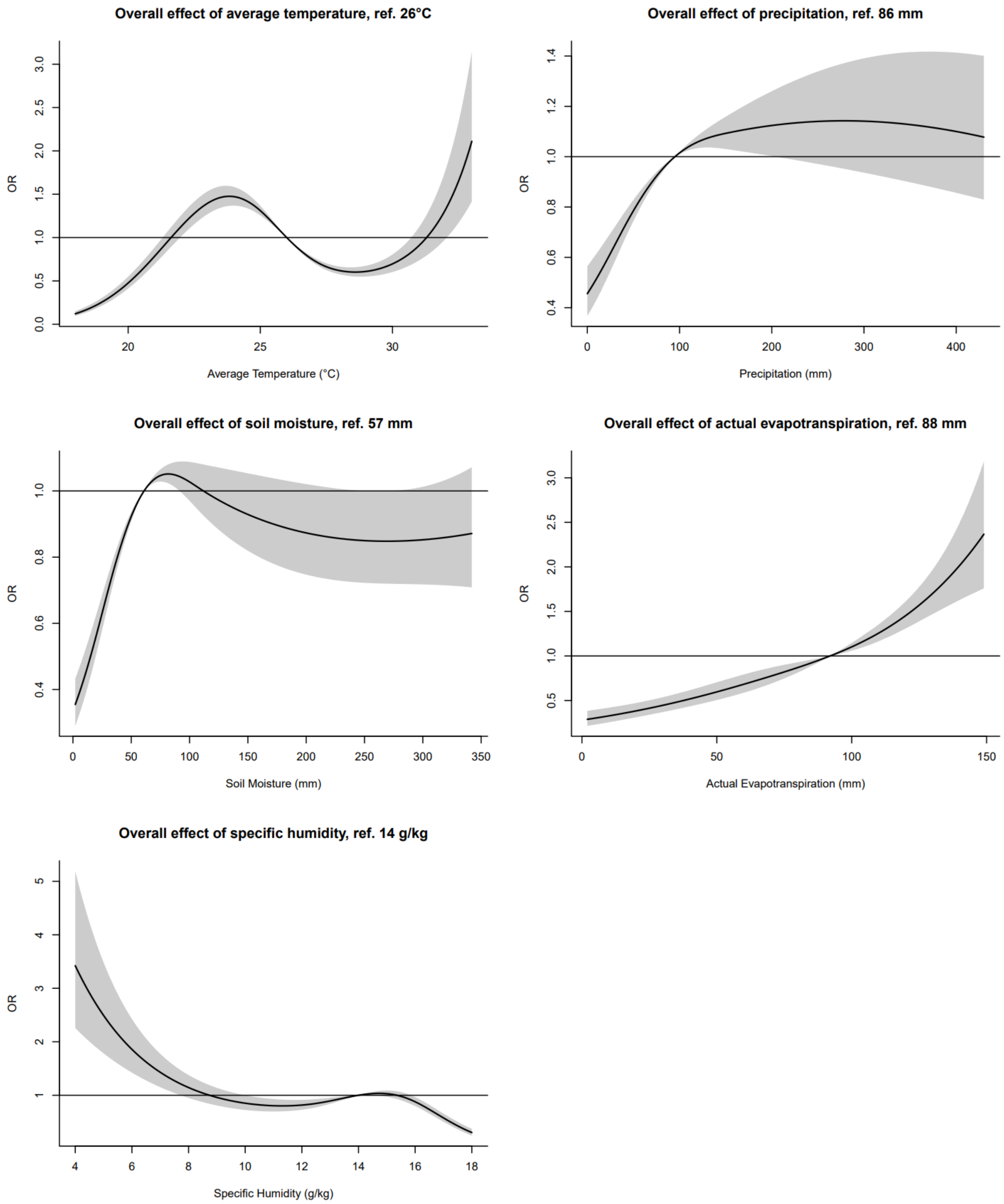

*Figure S2: Sensitivity analysis considering just microscopy tests.*

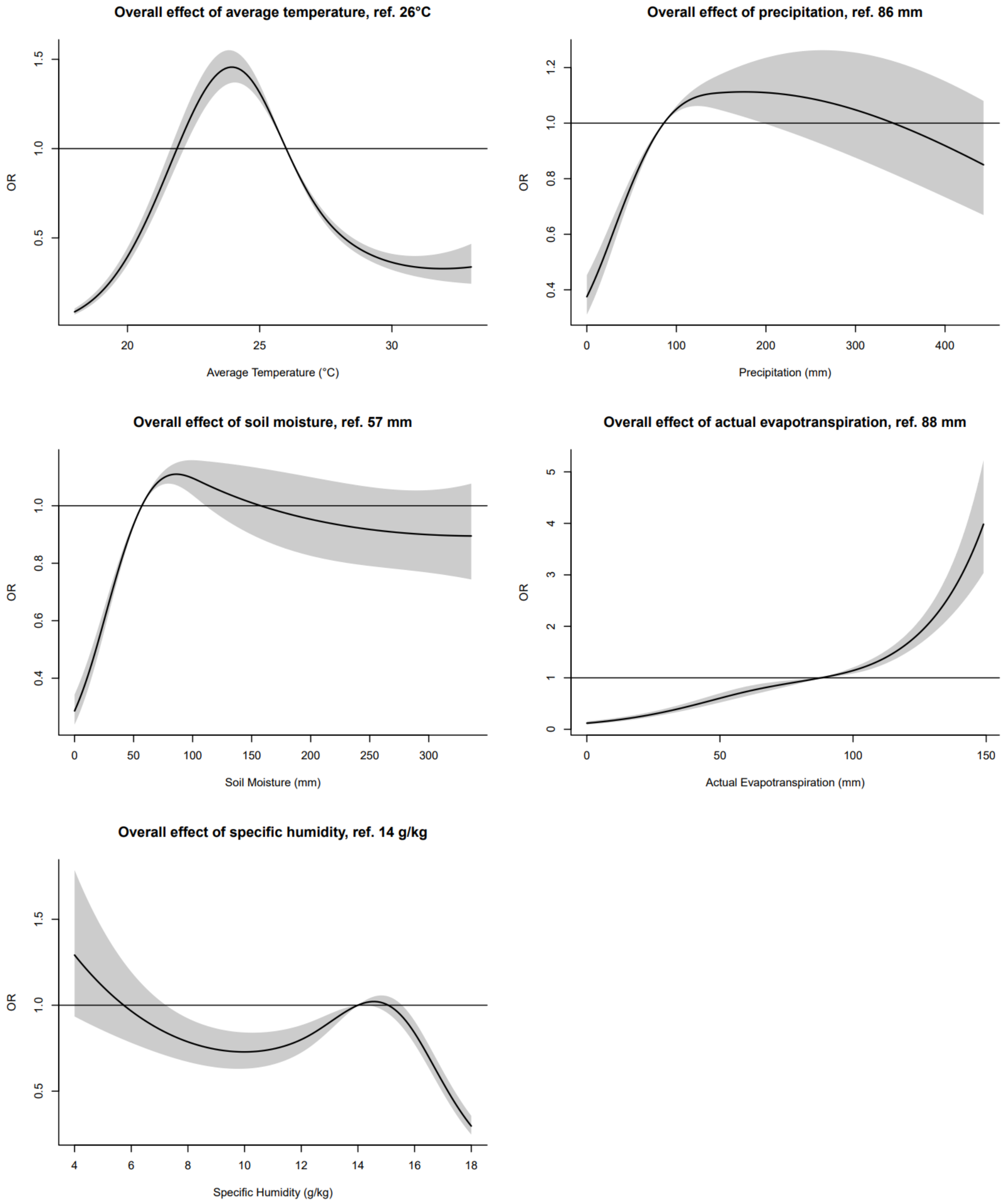

*Figure S3: Sensitivity analysis controlling for all lags of climatic exposures.*

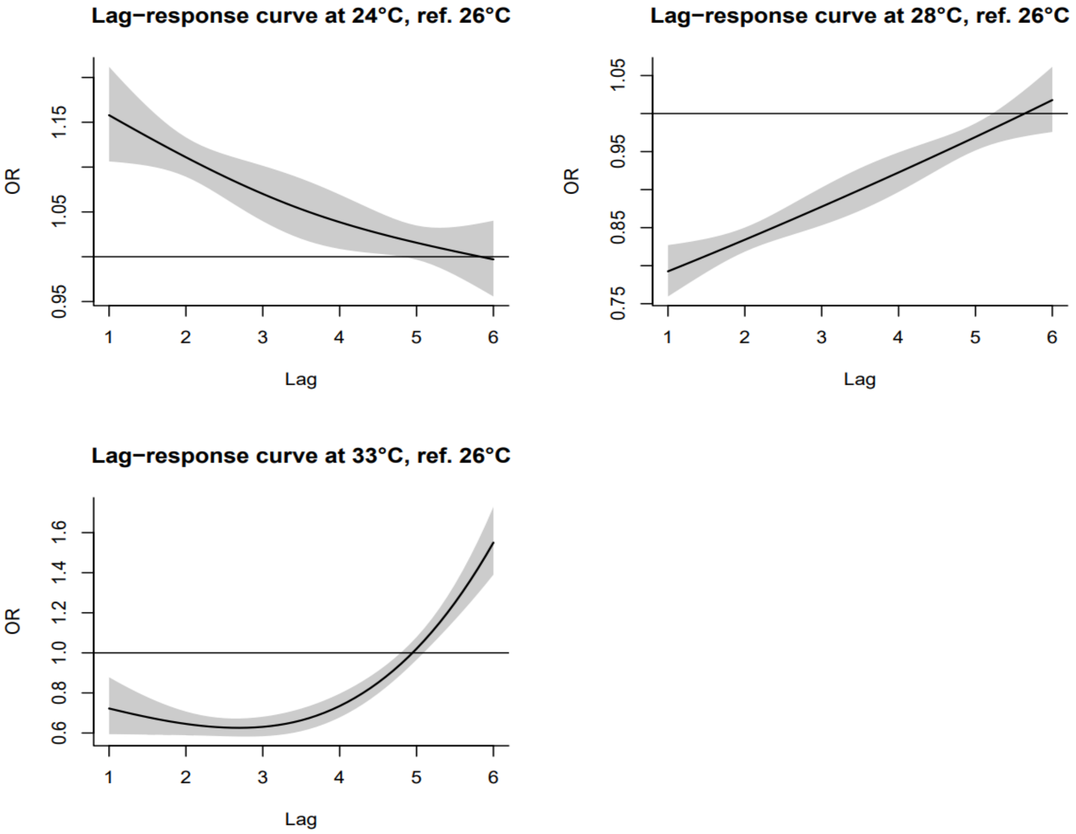

*Figure S4: Lag-response relationship at specific values of temperatures.*

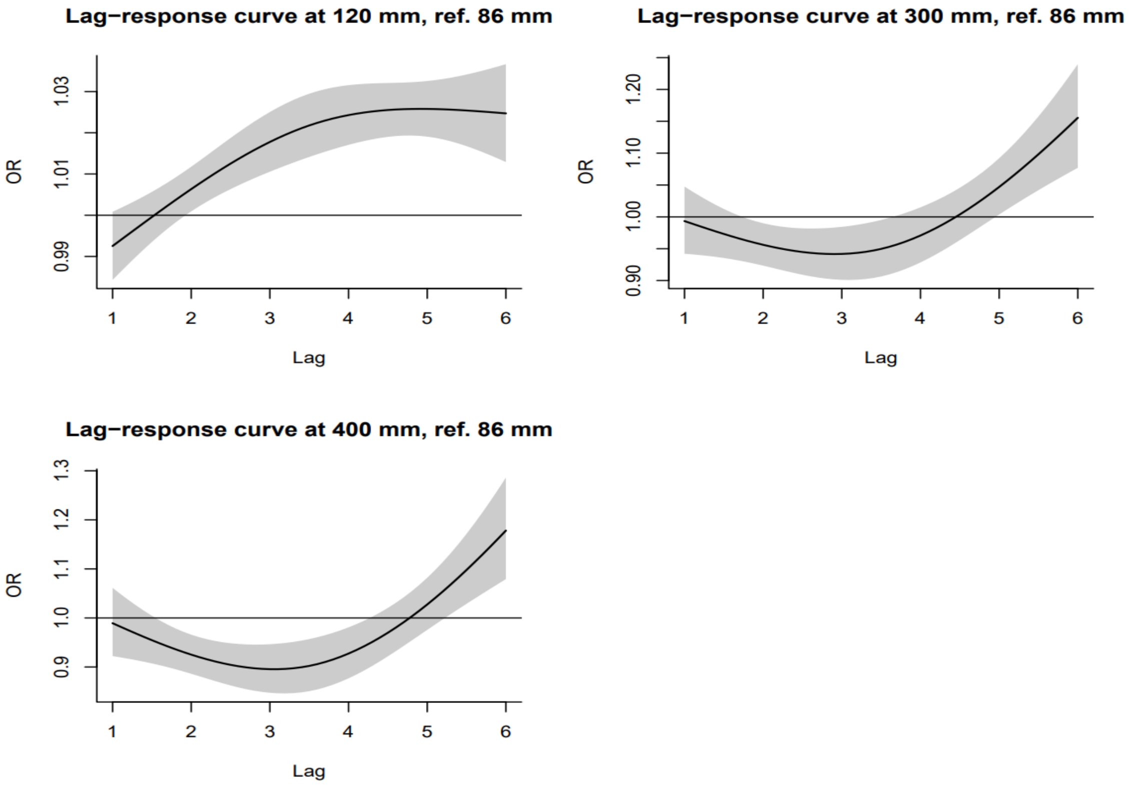

*Figure S5: Lag-response relationship at specific values of precipitation accumulation.*

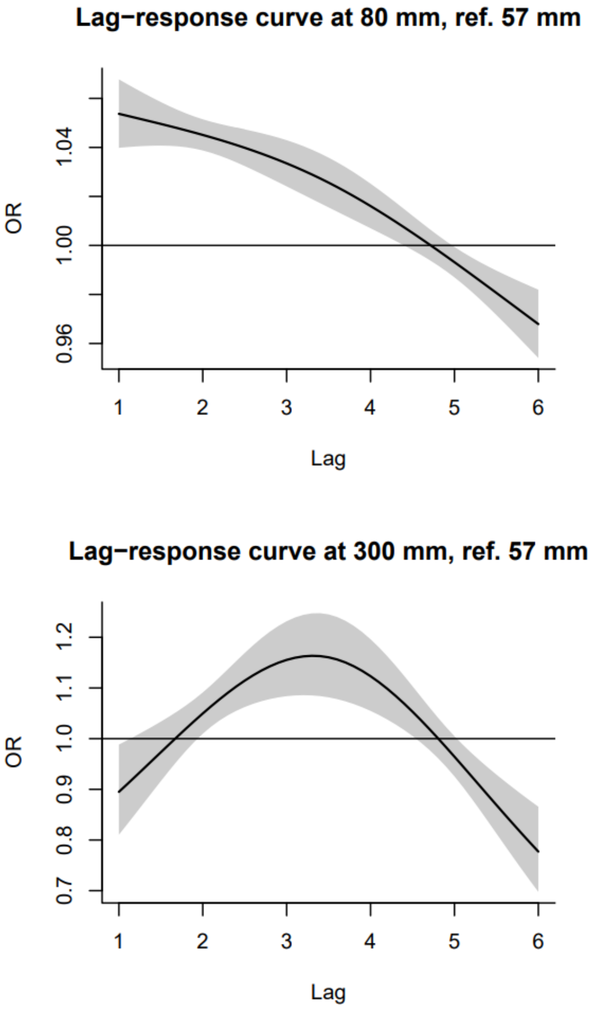

*Figure S6: Lag-response relationship at specific values of soil moisture.*

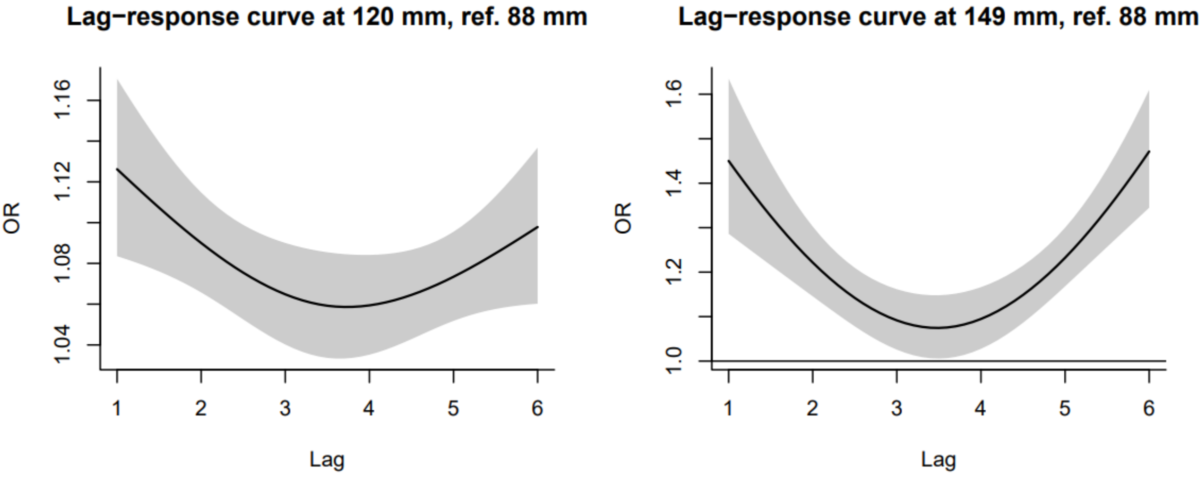

*Figure S7: Lag-response relationship at specific values of actual evapotranspiration.*

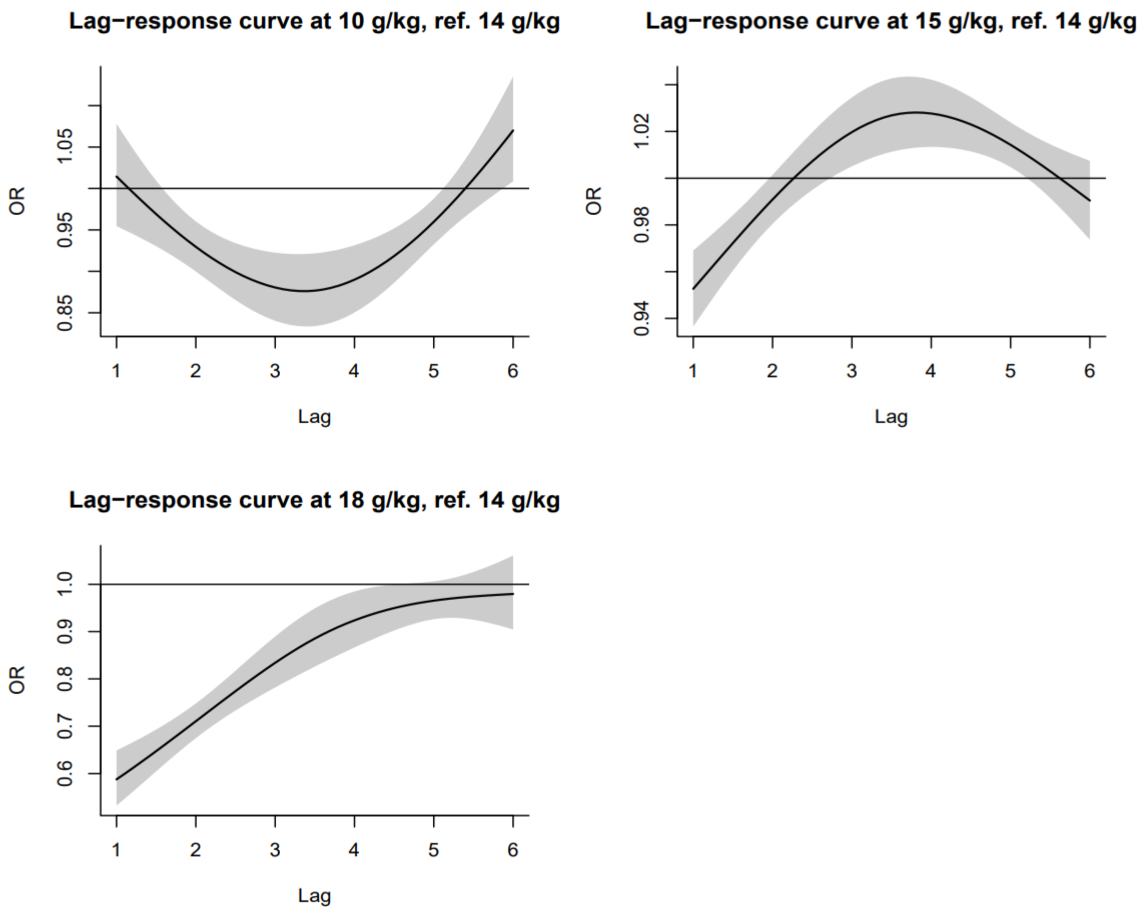

*Figure S8: Lag-response relationship at specific values of specific humidity.*

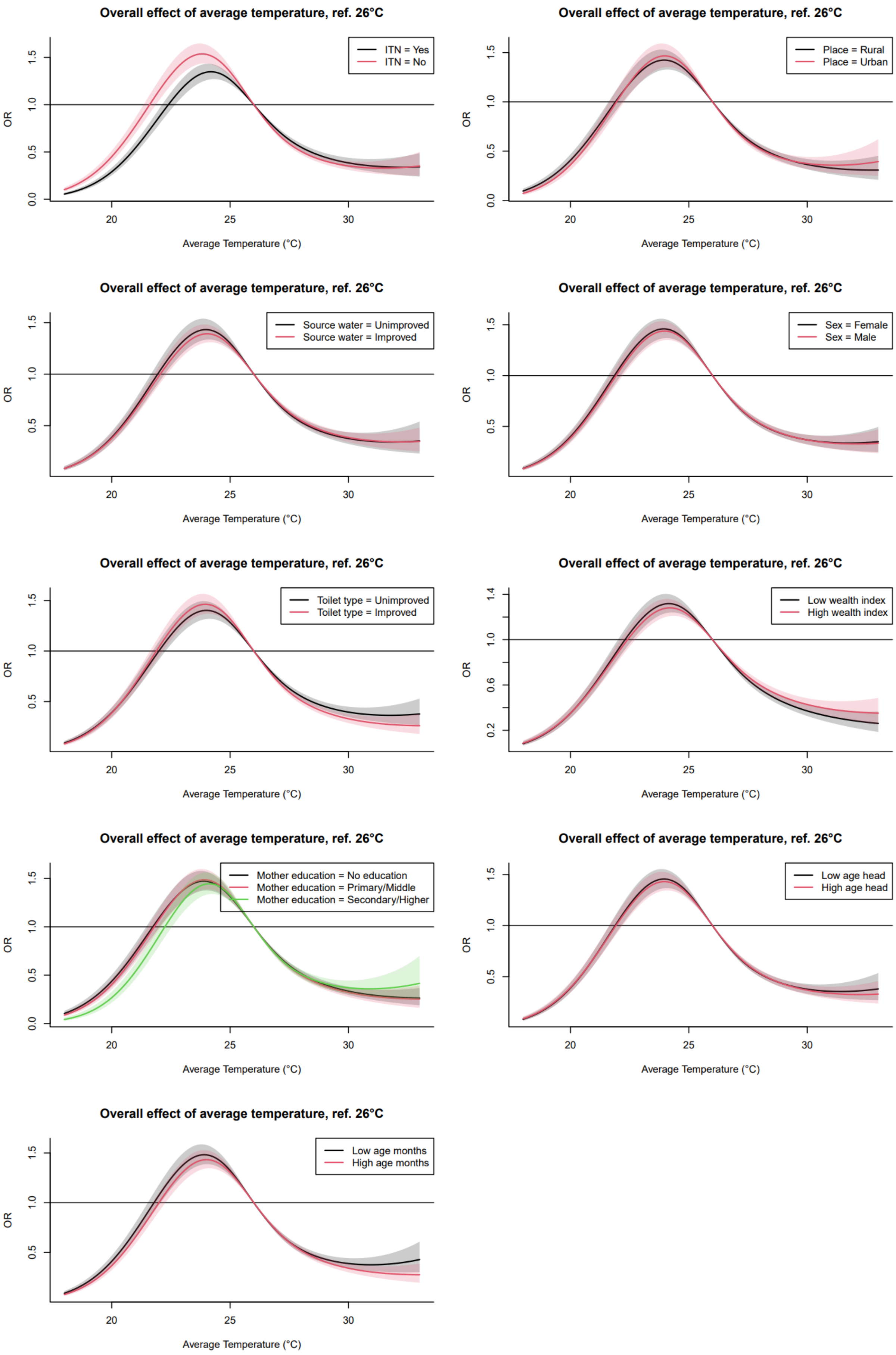

Figure S9: Effect modifiers for Average Temperature.

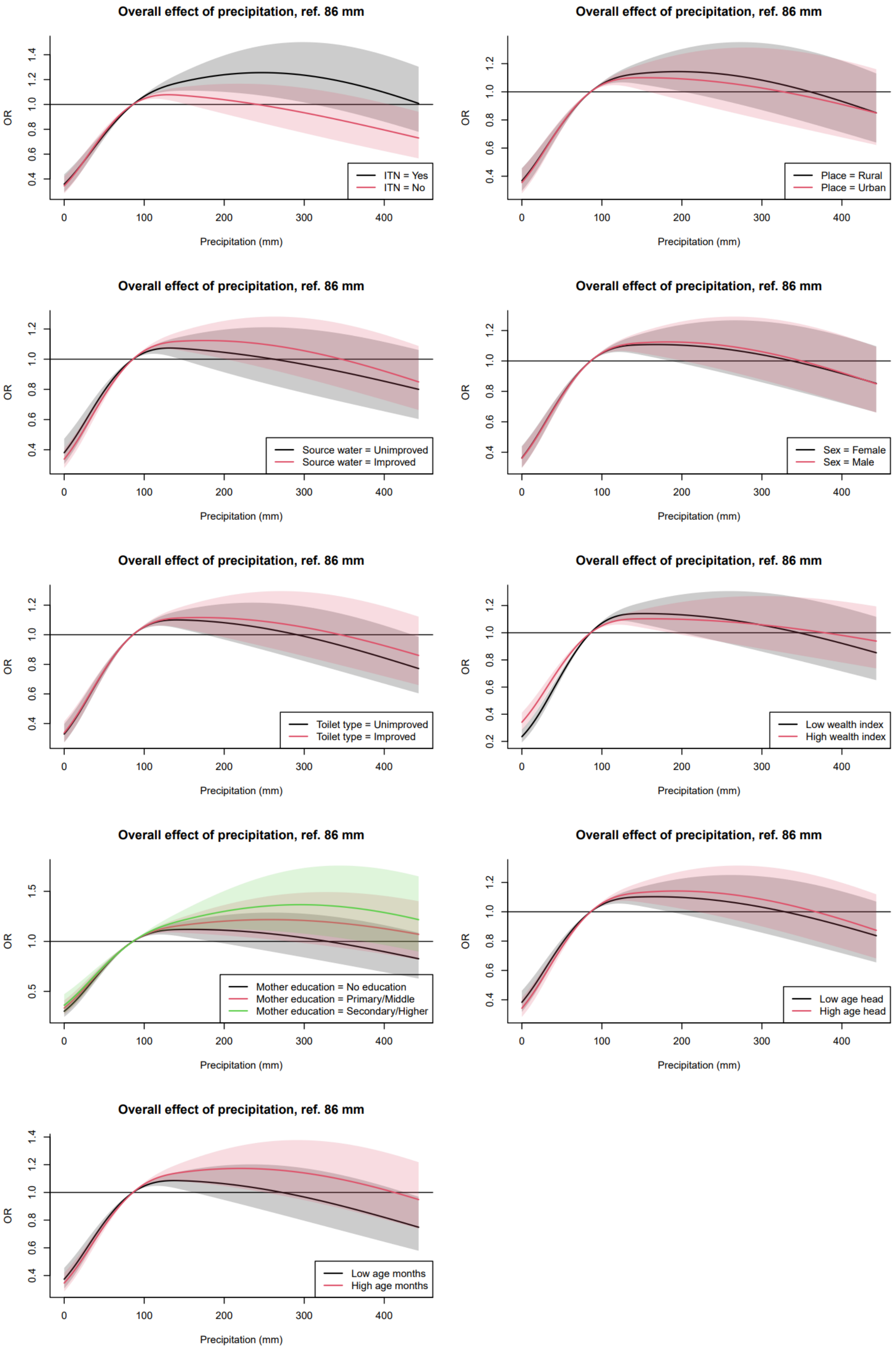

*Figure S10: Effect modifiers for Precipitation Accumulation.*

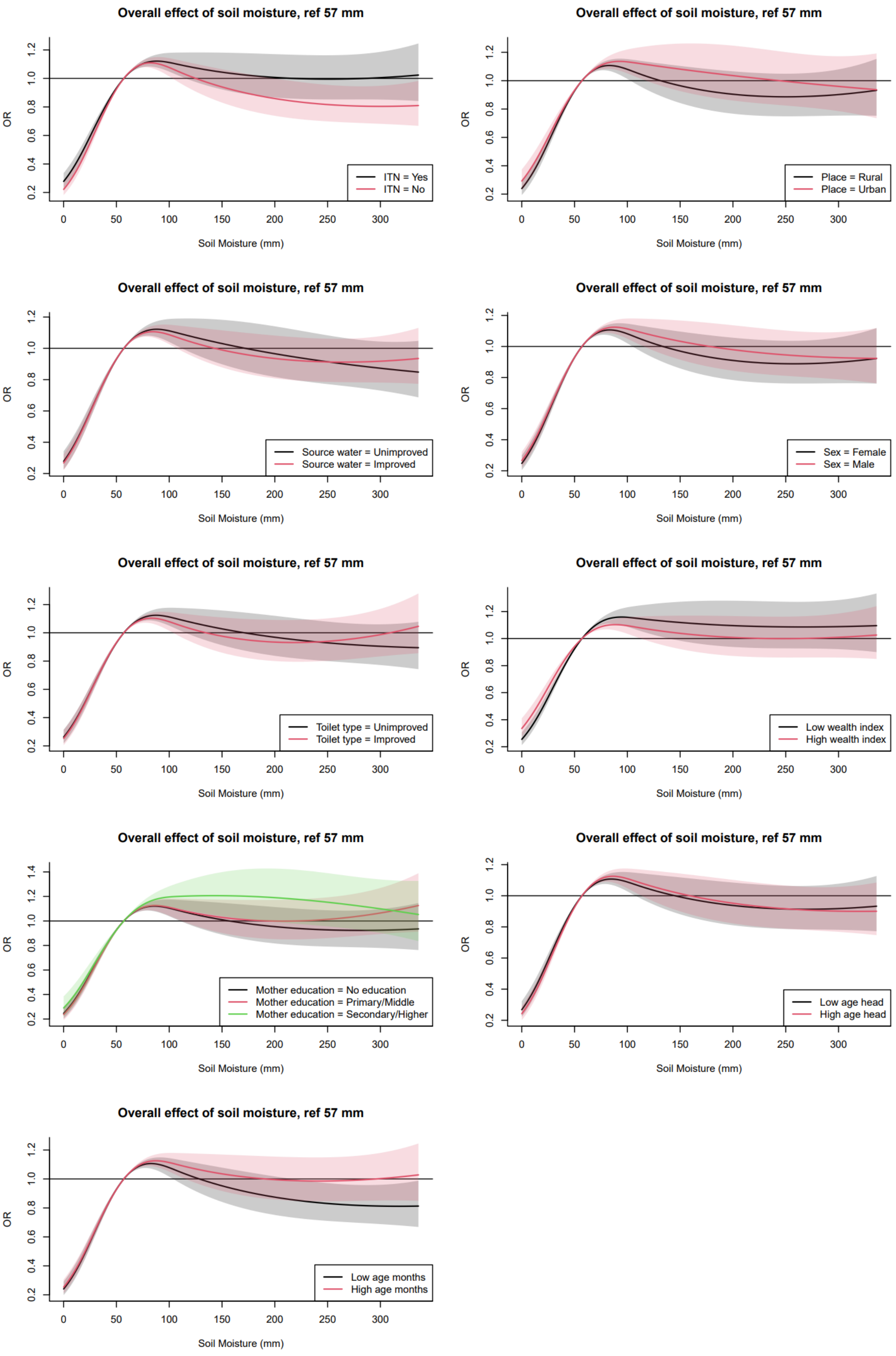

*Figure S11: Effect modifiers for Soil Moisture.*

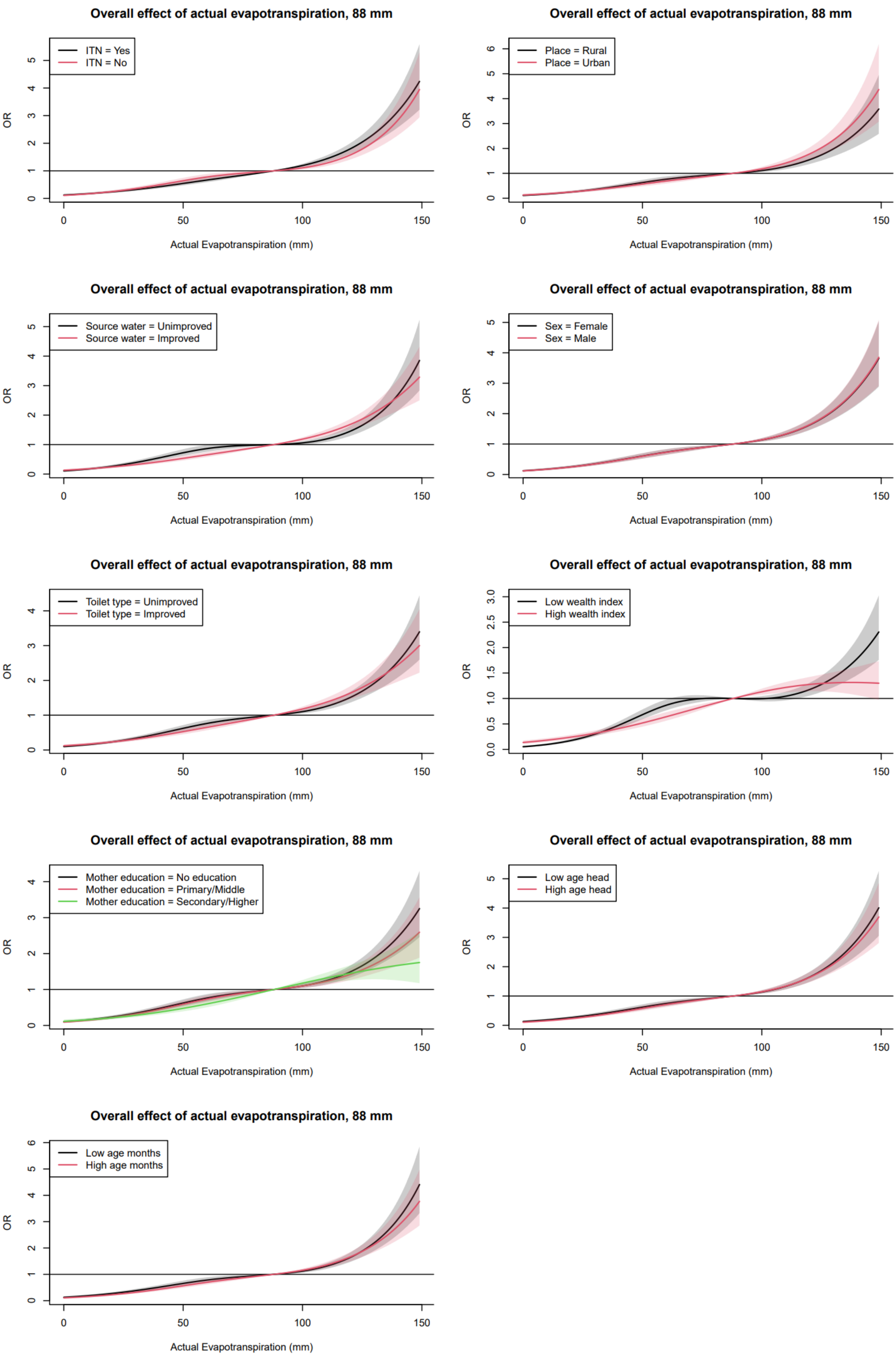

*Figure S12: Effect modifiers for Actual Evapotranspiration*

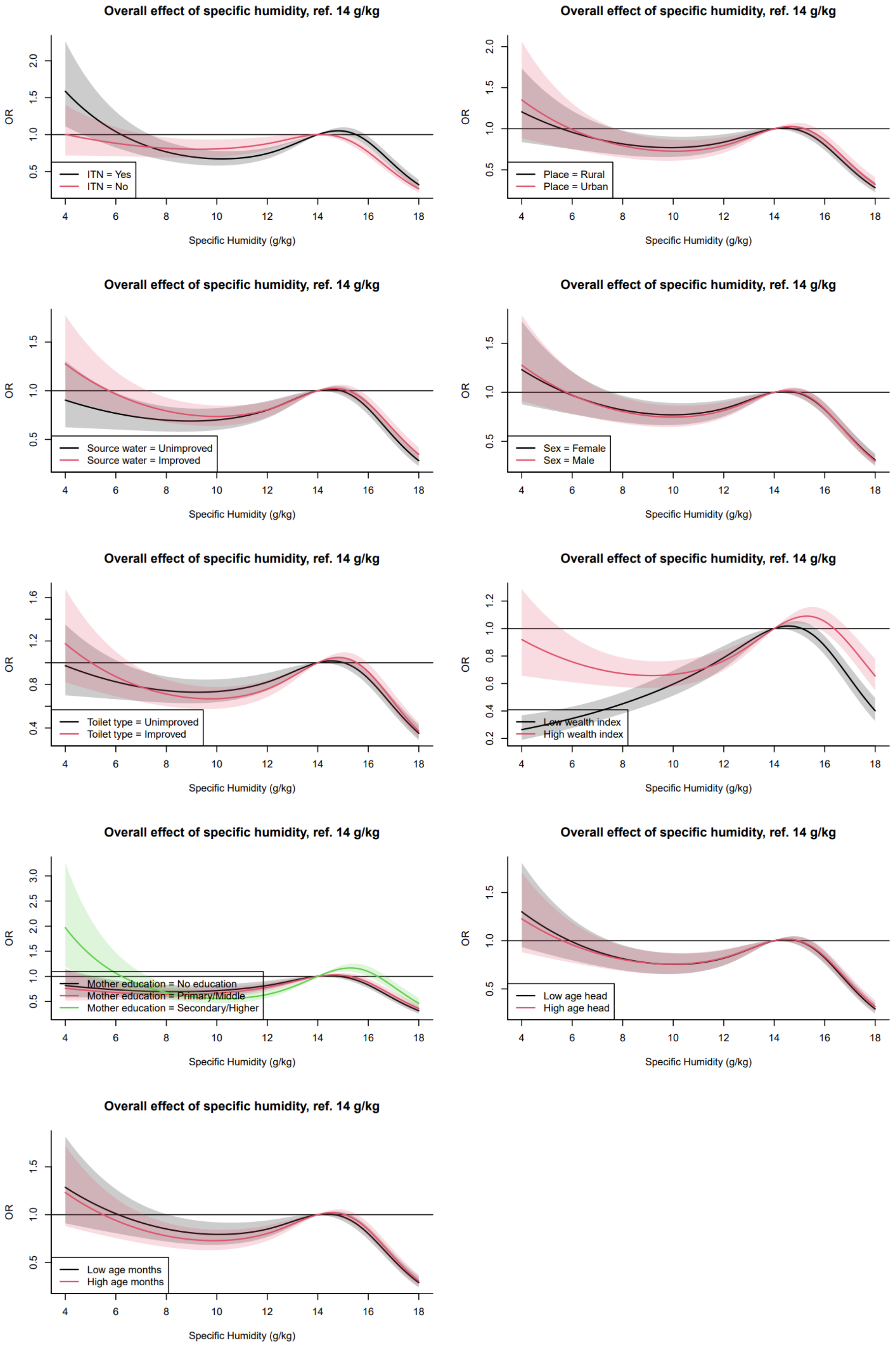

*Figure S13: Effect modifiers for Specific Humidity.*

#### *Supplementary Tables*

| **Exposure** | **Control Variables** |
|---|---|
| Temperature | Precipitation. |
| Precipitation | Temperature. |
| Soil Moisture | Temperature and Precipitation. |
| Evapotranspiration | Temperature, Precipitation and Soil Moisture. |
| Humidity | Temperature, Precipitation, Soil Moisture and Evapotranspiration. |

*Table S1: Control variables for each model.*

| | 2006 | 2007 | 2008 | 2009 | 2010 | 2011 | 2012 | 2013 | 2014 | 2015 | 2016 | 2017 | 2018 | 2019 | 2020 | 2021 | 2022 | 2023 |
|---|---|---|---|---|---|---|---|---|---|---|---|---|---|---|---|---|---|---|
| Angola | 1163 | 1147 | 0 | 0 | 0 | 3426 | 0 | 0 | 0 | 3224 | 3507 | 0 | 0 | 0 | 0 | 0 | 0 | 0 |
| Benin | 0 | 0 | 0 | 0 | 0 | 377 | 3714 | 0 | 0 | 0 | 0 | 3248 | 3151 | 0 | 0 | 0 | 0 | 0 |
| Burkina Faso | 0 | 0 | 0 | 0 | 6402 | 0 | 0 | 0 | 6242 | 0 | 0 | 554 | 4937 | 0 | 0 | 5688 | 0 | 0 |
| Burundi | 0 | 0 | 0 | 0 | 0 | 0 | 3733 | 5 | 0 | 0 | 3347 | 2341 | 0 | 0 | 0 | 0 | 0 | 0 |
| Cameroon | 0 | 0 | 0 | 0 | 0 | 0 | 0 | 0 | 0 | 0 | 0 | 0 | 4693 | 23 | 0 | 0 | 4344 | 0 |
| Côte d'Ivoire | 0 | 0 | 0 | 0 | 0 | 320 | 3041 | 0 | 0 | 0 | 0 | 0 | 0 | 0 | 0 | 4953 | 0 | 0 |
| DRC | 0 | 0 | 0 | 0 | 0 | 0 | 0 | 6583 | 1868 | 0 | 0 | 0 | 0 | 0 | 0 | 0 | 0 | 0 |
| Gabon | 0 | 0 | 0 | 0 | 0 | 0 | 0 | 0 | 0 | 0 | 0 | 0 | 0 | 403 | 2544 | 2867 | 0 | 0 |
| Gambia | 0 | 0 | 0 | 0 | 0 | 0 | 0 | 0 | 0 | 0 | 0 | 0 | 0 | 536 | 3049 | 0 | 0 | 0 |
| Ghana | 0 | 0 | 0 | 0 | 0 | 0 | 0 | 0 | 2717 | 0 | 3076 | 0 | 0 | 2868 | 0 | 0 | 4067 | 456 |
| Guinea | 0 | 0 | 0 | 0 | 0 | 0 | 3198 | 0 | 0 | 0 | 0 | 0 | 0 | 0 | 0 | 4052 | 0 | 0 |
| Kenya | 0 | 0 | 0 | 0 | 0 | 0 | 0 | 0 | 0 | 3271 | 0 | 0 | 0 | 0 | 3692 | 0 | 0 | 0 |
| Liberia | 0 | 0 | 498 | 3549 | 0 | 3185 | 0 | 0 | 0 | 0 | 2765 | 0 | 0 | 0 | 0 | 0 | 2924 | 0 |
| Madagascar | 0 | 0 | 0 | 0 | 0 | 6122 | 0 | 5340 | 0 | 0 | 6743 | 0 | 0 | 0 | 0 | 5866 | 0 | 0 |
| Malawi | 0 | 0 | 0 | 0 | 0 | 0 | 2078 | 0 | 1949 | 0 | 0 | 2310 | 0 | 0 | 0 | 0 | 0 | 0 |
| Mali | 0 | 0 | 0 | 0 | 0 | 0 | 3091 | 1799 | 0 | 7198 | 0 | 0 | 4257 | 0 | 0 | 8870 | 0 | 0 |
| Mauritania | 0 | 0 | 0 | 0 | 0 | 0 | 0 | 0 | 0 | 0 | 0 | 0 | 0 | 2294 | 6524 | 1048 | 0 | 0 |
| Mozambique | 0 | 0 | 0 | 0 | 0 | 4778 | 0 | 0 | 0 | 0 | 0 | 0 | 4286 | 0 | 0 | 0 | 3537 | 706 |
| Niger | 0 | 0 | 0 | 0 | 0 | 0 | 0 | 0 | 0 | 0 | 0 | 0 | 0 | 0 | 0 | 4870 | 0 | 0 |
| Nigeria | 0 | 0 | 0 | 0 | 5087 | 0 | 0 | 0 | 0 | 5966 | 0 | 0 | 10727 | 0 | 0 | 11004 | 0 | 0 |
| Rwanda | 0 | 0 | 0 | 0 | 2293 | 1764 | 0 | 0 | 1247 | 2283 | 0 | 0 | 0 | 1011 | 2674 | 0 | 0 | 0 |
| Senegal | 0 | 0 | 0 | 0 | 1491 | 2376 | 2147 | 3836 | 6153 | 6150 | 6011 | 10722 | 0 | 0 | 513 | 3778 | 0 | 0 |
| Sierra Leone | 0 | 0 | 0 | 0 | 0 | 0 | 0 | 0 | 0 | 0 | 6536 | 0 | 0 | 0 | 0 | 0 | 0 | 0 |
| Tanzania | 0 | 0 | 0 | 0 | 0 | 0 | 0 | 0 | 0 | 6781 | 2522 | 7167 | 0 | 0 | 0 | 0 | 5168 | 0 |
| Togo | 0 | 0 | 0 | 0 | 0 | 0 | 0 | 1172 | 2041 | 0 | 0 | 3209 | 0 | 0 | 0 | 0 | 0 | 0 |
| Uganda | 0 | 0 | 0 | 3683 | 11 | 0 | 0 | 0 | 2876 | 1686 | 4641 | 0 | 2514 | 4431 | 0 | 0 | 0 | 0 |

*Table S2: Distribution of the sample over country and year.*

| **Effect modifier** | **Category** | **Odds Ratio** | **p-value** |
|---|---|---|---|
| ITN net | *No* | Ref | |
| | *Yes* | 0.84 | <0.001 |
| Type of Residency | *Rural* | Ref | |
| | *Urban* | 0.94 | 0.006 |
| Household source of drinking water | *Unimproved* | Ref | |
| | *Improved* | 0.83 | <0.001 |
| Sex of the child | *Female* | Ref | |
| | *Male* | 1.03 | 0.004 |
| Household type of toilet facility | *Unimproved* | Ref | |
| | *Improved* | 0.76 | <0.001 |
| Mother's education level | *No education* | Ref | |
| | *Primary/Middle* | 0.88 | <0.001 |
| | *Secondary/Higher* | 0.59 | <0.001 |
| | *Missing* | 0.99 | 0.813 |
| Relative Wealth Index (RWI) | | 0.36 | <0.001 |
| Age of head of household | | 1.00 | 0.031 |
| Age of the child | | 1.02 | <0.001 |

*Table S3: Main effects of effect modifiers controlling for the climatic variables.*

| **Effect Modifier** | **Temperature** | **Precipitation** | **Soil Moisture** | **Evapotranspiration** | **Humidity** |
|---|---|---|---|---|---|
| ITN net | <0.001*** | <0.001*** | <0.001*** | <0.001*** | <0.001*** |
| Type of residency | 0.043* | <0.001*** | 0.002** | <0.001*** | <0.001*** |
| Household source of drinking water | 0.002** | <0.001*** | 0.004** | <0.001*** | <0.001*** |
| Sex of the child | 0.702 | 0.968 | 0.663 | 0.889 | 0.985 |
| Household type of toilet facility | <0.001*** | <0.001*** | <0.001*** | <0.001*** | <0.001*** |
| Relative Wealth Index (RWI) | <0.001*** | <0.001*** | <0.001*** | <0.001*** | <0.001*** |
| Mother's educational level | <0.001*** | 0.003** | <0.001*** | <0.001*** | <0.001*** |
| Age of head of household | 0.002** | 0.004* | <0.001*** | <0.001*** | 0.001** |
| Age of the child | <0.001*** | <0.001*** | <0.001*** | <0.001*** | <0.001*** |

*Table S4: p-values Likelihood Ratio Tests (LRT) of interaction terms for all climatic exposures.*
